# Improved Sensitivity of Digits-in-Noise Test to High-Frequency Hearing Loss

**DOI:** 10.1101/2020.07.31.20165225

**Authors:** Lina Motlagh Zadeh, Noah H. Silbert, De Wet Swanepoel, David R. Moore

## Abstract

**Objectives:** Hearing loss is most commonly observed at high frequencies. High-frequency hearing loss (HFHL) precedes and predicts hearing loss at lower frequencies. It was previously shown that an automated, self-administered digits-in-noise (DIN) test can be sensitized for detection of HFHL by low-pass filtering the speech-shaped masking noise at 1.5 kHz. This study was designed to investigate whether sensitivity of the DIN to HFHL can be enhanced further using low-pass noise filters with higher cutoff frequencies.

**Design:** US-English digits 0-9, homogenized for audibility, were binaurally presented in different noise maskers including one broadband and three low-pass (cut-off at 2, 4, 8 kHz) filtered speech-shaped noises. DIN-Speech reception thresholds (SRTs) were obtained from 60 normal hearing (NH), and 40 mildly hearing impaired (HI) listeners with bilateral symmetric sensorineural hearing-loss. Standard and extended high frequency audiometric pure tone averages (PTAs) were compared with the DIN-SRTs.

**Results:** Narrower masking noise bandwidth generally produced better (more sensitive) mean DIN-SRTs. There were strong and significant correlations between SRT and PTA in the HI group. Lower frequency, PTA_LF 0.5,1, 2, 4 kHz_ had the highest correlation and steepest slope with SRTs obtained from the 2 kHz filter. Higher frequency, PTA_HF 4,8,10,12.5 kHz_ correlated best with SRTs obtained from 4 and 8 kHz filtered noise. The 4 kHz low-pass filter also had the highest sensitivity (92%) and equally-highest (with the 8 kHz filter) specificity (90%) for detecting an average PTA_HF_ of 20 dB or more.

**Conclusions:** Of the filters used, DIN sensitivity to higher frequency hearing loss was greatest using the 4 kHz low-pass filter. These results suggest that low-pass filtered noise may be usefully substituted for broadband noise to improve earlier detection of HFHL using DIN.

## INTRODUCTION

Late-diagnosed hearing loss in the elderly leads to multiple negative consequences, including poor quality of life, depression, social isolation, and other psychosocial disorders (Chia et al. 2007; Stika and Hays 2015). The annual global cost of unaddressed hearing loss is estimated at 750 billion dollars (WHO, 2019). Early detection and treatment of hearing loss is both care- and cost-effective in preserving hearing and quality of life for affected people (Davis et al. 2007; Karpa et al. 2010). Hearing screening plays a key role in early diagnosis of hearing loss (Davis et al., 2007, Chia et al. 2007). Development and validation of an accurate, accessible, inexpensive, fast, and objective hearing screening that can reliably detect early signs of hearing loss is expected to provide significant benefits, especially for populations who do not have ready access to audiology services (Leensen et al. 2011; Potgieter et al. 2016; Swanepoel et al. 2019)

### Detection of high-frequency hearing loss (HFHL)

High frequencies are the first part of the hearing spectrum to be lost in most forms of hearing loss (Dubno et al. 2013). The most common contributing factors that cause HFHL in adults include age, noise exposure, and ototoxic drugs (Gratton and Vázquez 2003; Mehrparvar et al, 2011; Vlaming et al, 2014; Yang et al, 2015). Age-related hearing loss (presbycusis) is thought to begin with degeneration of outer hair cells in the basal end of the cochlea, causing a HFHL that gradually affects lower frequencies (Gratton and Vázquez 2003). Noise-induced hearing loss (NIHL) is a result of prolonged exposure to loud sounds and also mainly affects high frequencies (Le et al. 2017; Mehrparvar et al. 2011). Both presbycusis and NIHL can negatively influence speech perception (Liberman 2017); the majority of meaning within speech is delivered by consonants (Nespor et al. 2003), the discrimination of some of which is strongly influenced by high frequency signals (Vitela et al. 2015; Patel et al. 2018). Therefore, developing a reliable hearing screening test with high sensitivity to HFHL is desirable for detection and possible early prevention of later disabling hearing loss.

Most studies of HFHL and NIHL have used pure tone audiometry as the sole basis of measurement. However, pure tone audiometry may not be the best, or should not be the only, predictor of the difficulty a person will have listening to speech in a challenging environment (Killion and Niquette 2000; Vermiglio et al. 2012; Moore et al. 2014; Moore et al. 2017). Speech signals have a spectro-temporal complexity that changes with a limited predictability over time and accurate speech coding and recognition requires multiple auditory discrimination skills (Summers et al. 2013). Pure tone detection only requires minimal cognitive resources, in contrast to supra-threshold speech perception, especially in the presence of competing noise (Moore et al. 2014). Digits-in-noise (DIN) is a relatively undemanding speech-in-noise test that allows us to objectively, reliably and quickly measure speech recognition abilities in addition to retaining a close relationship to audiometry (Smits et al. 2004; Smits et al. 2006; Jansen et al. 2010; Vlaming et al. 2014).

Previous studies have shown the utility of DIN as an objective and reliable hearing screen (Smits et al. 2006; Ozimek et al. 2009; Leensen et al. 2011; Vlaming et al. 2014; Folmer et al. 2017). The DIN typically presents digits to listeners against a simultaneous background of broad-bandwidth, speech-spectrum-shaped noise. A ‘speech reception threshold’ (SRT) is defined as the speech (digit) signal-to-noise ratio at which three successive digits are all correctly recognized on 50% of presentations (Vlaming et al. 2014). DIN screening tests have been developed in several languages (English, Danish, French, German, etc.) and are available in self-administered forms, deliverable via telephone (Smits et al. 2004), computer (Folmer et al. 2017) or smartphone/tablet (Potgieter et al. 2016; De Sousa et al. 2018) in any moderately quiet setting. Studies have reported high levels of DIN sensitivity and specificity, and a strong correlation between DIN-SRTs and audiometric ‘pure tone average’ (PTA) measures in adult listeners of mixed hearing ability (Jansen et al. 2010; Smits et al. 2013; Vlaming et al. 2014).

Digits (0-9) have been used as stimuli in several speech perception tests (Ramkissoon et al. 2002; Wilson et al. 2005) and in clinical screening (Smits and Houtgast 2007; Ozimek et al. 2009; Smits et al. 2013; Vlaming et al. 2014). Digits are highly overlearned stimuli that are easily recognized by a wide range of people including young children (Koopmans et al. 2018) and, especially in the case of English digits, by non-native language speakers (Smits et al. 2016). Randomized digit triplets have a low probability of being guessed and provide more accurate and reproducible SRT estimates than other speech test materials, including non-words, non-digit words, single digits, and sentences (Jansen et al. 2013; Smits et al. 2013; Vlaming et al. 2014).

During the past 10 years, different versions of English telephone- and computer-based DIN tests have been developed for detection of impaired hearing (Smits et al. 2006; Leensen et al. 2011; Folmer et al. 2017). However, there has been concern that, since English digits may be largely distinguished based on their low-frequency vowel formants, the DIN could be insensitive to HFHL (Smits et al. 2016). To address this concern, Vlaming and colleagues (2014) used low-pass noise filtering (1.5 kHz cutoff) to increase the relative audibility of the higher frequency components of the speech signals and thus to improve the sensitivity of the DIN to HFHL (in this case, PTA_3,4,6,8 kHz_). They found higher sensitivity and specificity (87% and 92% respectively) for this ‘high frequency DIN’ compared to the standard DIN that uses broadband noise masking. Surprisingly, they also found higher sensitivity of the high frequency DIN than the standard, broadband masked DIN to lower tone frequencies (PTA_0.5,1,2,4 kHz_). One possible explanation for this finding could be the higher degree of hearing loss at 2 and 4 kHz in most of those with low-frequency hearing loss.

The maximum frequency that was tested audiometrically by Vlaming and colleagues (2014) was 8 kHz. Although frequencies below 6 kHz provide most of the required phonetic information for speech perception, substantial evidence suggests that there is salient information that affects speech intelligibility in the extended high frequency (EHF; 10-16 kHz) regions (Knight et al. 2007; Le Prell et al. 2013; Vitela et al. 2015; Rodríguez Valiente et al. 2016). In a recent study, Motlagh Zadeh et al. (2019) showed the considerable contribution of EHF energy to speech perception in noise. In that study, two thirds (74/116) of a sample of mostly younger adults with normal, conventional frequency (0.25-8 kHz) audiograms had EHF hearing loss. Interestingly, about the same two-thirds reported difficulty hearing in an everyday noisy situation. EHF hearing loss was positively related to the number of individuals self-reporting difficulty in that situation, suggesting that widespread EHF hearing loss among those with normal standard audiograms may contribute to the common report of difficulty hearing in noisy places. This study aimed to investigate whether sensitivity of the DIN to HFHL can be enhanced further using higher cutoff frequency (2, 4, and 8 kHz) low-pass noise filters. We hypothesized, first, that the higher the cut-off frequency of the filtered noise, the more sensitive the test to HFHL and, second, that hearing screening sensitive to a broader range of HFHL (to 16 kHz) may further sensitize the test, potentially aiding detection of the earliest signs of hearing loss.

## MATERIALS AND METHODS

### Participants

In a previously reported study (Motlagh-Zadeh et al. 2019), we recruited 70 people (44 female, Mean = 29.4 y/o, SD = 10.2) via advertisements sent to staff at Cincinnati Children’s Hospital Medical Center (CCHMC), and flyers distributed in the community. Pure-tone audiometry was performed from 0.25-16 kHz using an Interacoustics Equinox 2.0 model audiometer calibrated to ANSI 3.6 (2010). Participants were tested in a double-walled sound booth (Acoustic Systems, Austin, Texas) meeting criteria of ANSI S3.1-1999 for audiometric test rooms. Air conduction thresholds were obtained using Sennheiser HDA300 circumaural headphones for all test frequencies. All these participants had normal hearing sensitivity (≤ 20 dB HL) in both ears across the range of audiometric frequencies from 0.25-8 kHz. Ten listeners performed homogenization calibration for the DIN to equalize signal/noise ratio for each digit at 50 % intelligibility, and 60 participated in the validation part of the study.

Using the same recruitment methods, an additional 83 people responded to an advertisement for this study that invited participation of those who had a diagnosis of sensorineural hearing loss, or were suspected of having a hearing loss. However, following the same audiometric procedure described above, only 40 of these people (average age = 54.2, SD = 9.2, 19 female) met the inclusion criteria for hearing impairment (HI; > 20 dB HL sensorineural hearing loss in each ear, confirmed with bone conduction audiometry, at any frequency from 0.25-8 kHz, and no more than 20 dB interaural asymmetry). All participants gave written informed consent and were paid under the approval of the CCHMC Institutional Review Board. Figure 1 shows the mean (and SD) hearing thresholds for both normal-hearing (NH) and hearing-impaired (HI) groups. Compared with some studies (e.g. Vlaming et al., 2014), the HI group had relatively mild hearing losses, especially at lower frequencies.

**Figure 1.**
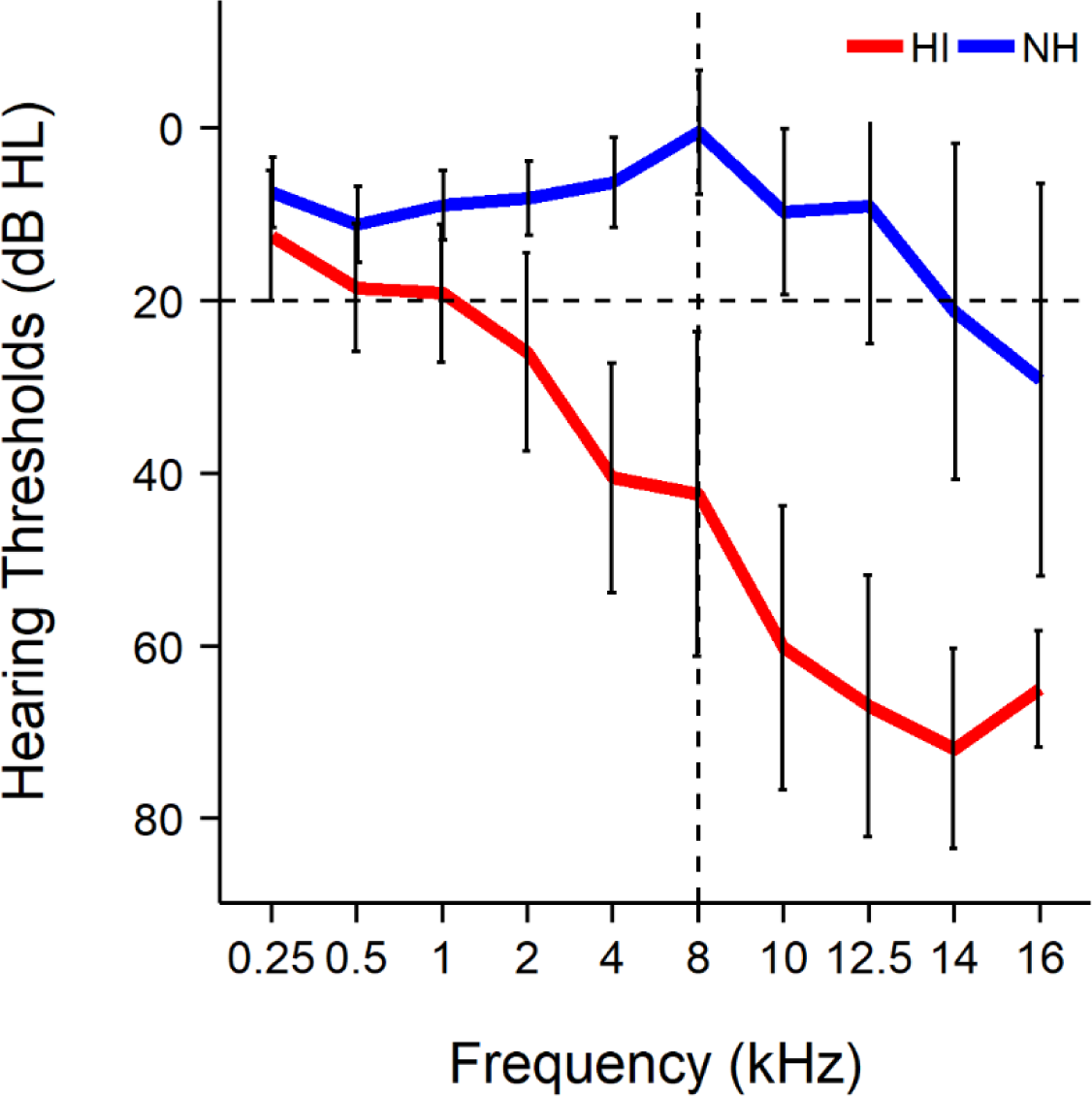
Binaural mean (± 1 SD) hearing thresholds for the normal-hearing (NH) and hearing-impaired (HI) groups. Black dashed horizontal line shows the level of normal hearing sensitivity (≤ 20 dB HL). Black dashed vertical line indicates standard range of the audiogram (0.25-8 kHz).

To provide further insight into the hearing ability and inter-listener variability of the HI group, Figure 2 shows the individuals with most and least hearing loss, and those closest to 25th and 75th percentile based on lower frequency, PTA_LF 0.5, 1, 2, 4 kHz_ and higher frequency, PTA_HF 4, 8,10, 12.5 kHz_. The PTAs referred to hearing thresholds of the better ear (whether left or right). Note that some participants with HI according to these criteria had PTA_LF_ and PTA_HF_ ≤ 20 dB HL.

**Figure 2.**
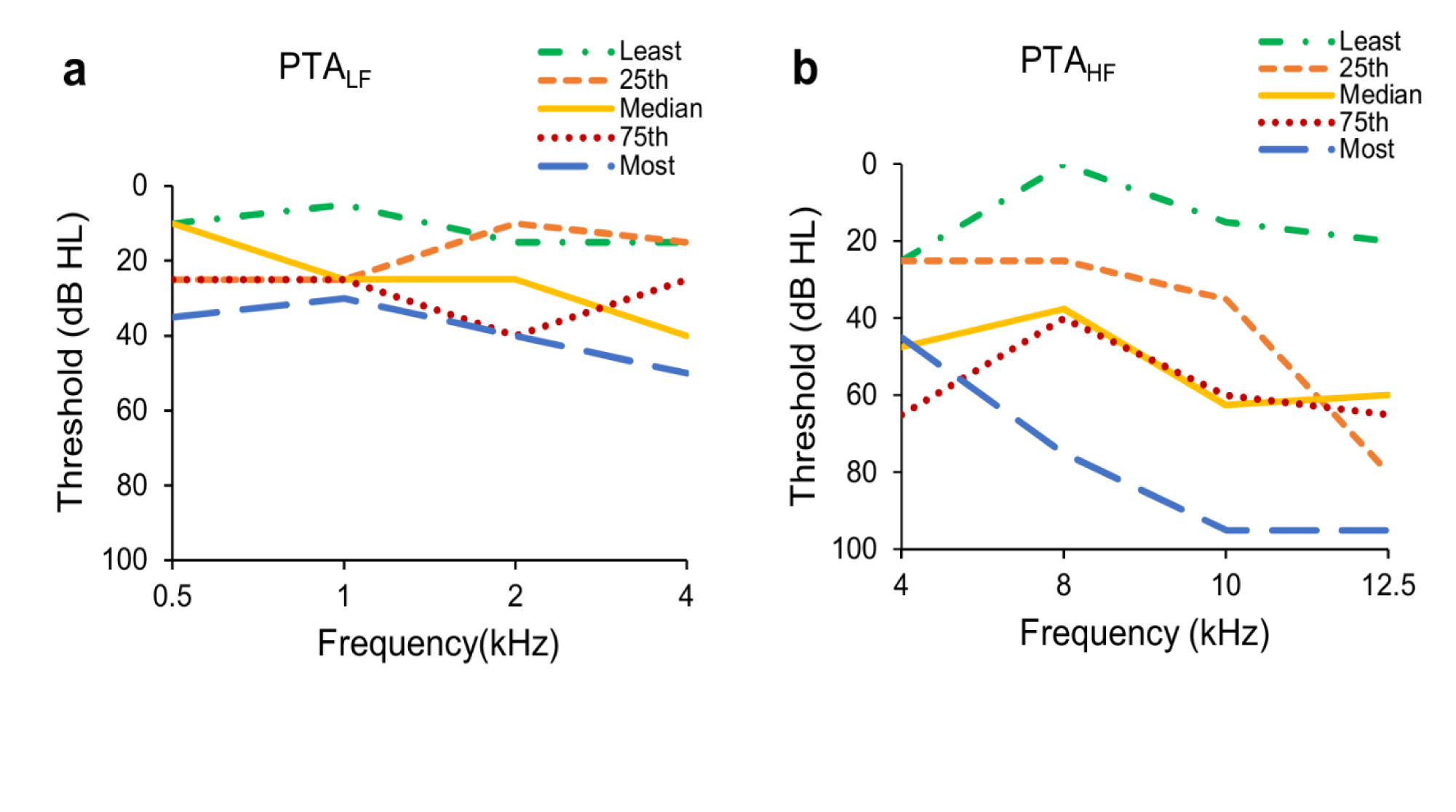
Hearing thresholds of the better ear for individual listeners in the hearing impaired (HI) group with the least and most hearing loss, and those closest to median, and 25^th^ and 75th percentile. Individuals were chosen based on audiometric pure-tone average (PTA) of (Panel **a**) lower (PTA_LF_ = PTA_0.5,1,2,4 kHz_), and (Panel **b**) higher (PTA_HF_ = PTA_4,8,10,12.5 kHz_) frequencies.

### Digits-in-noise test (DIN)

The DIN was developed based on the method described by Vlaming and colleagues (2014) and the digit level homogenization and validation processes have been reported in detail in Motlagh Zadeh et al. (2019).

### Stimuli

The DIN stimuli used in the present study were developed by Motlagh-Zadeh et al (2019). A list of 20 triplets was made from 10 digits (0 to 9, including bisyllabic ‘zero’ and ‘seven’; see Smits 2016) that were homogenized for equal intelligibility. The test triplets were made by connecting three different digits with an inter-digit interval of ∼175 ms. Triplets were presented diotically in one of 4 different noise maskers, one broadband (BB) and three low-pass filtered (cutoff at 2, 4, 8 kHz) speech-shaped noise. The BB masker was developed using the average frequency spectrum across all digits. Low-pass noise versions of the masker were constructed using a 10th-order Butterworth low-pass filter with three different cutoff frequencies (2, 4, 8 kHz), summed with a 15 dB attenuated version of the original broadband noise. Masking noise was started 100 ms before and ended 100 ms after each triplet presentation, and was interrupted in between successive triplets. For detailed information regarding recordings, homogenizationg, and noise maskers construction refer to the Supplementary Information link provided by Motlagh-Zadeh et al (2019).

### Procedures

SRTs were obtained using an up-down adaptive procedure in which, following each correct response, the SNR level was reduced (became more difficult) by 2 dB and, following each incorrect response, the SNR increased by 2 dB. All three digits of a triplet had to be correct to count as a correct response. Digit level roamed and noise level was fixed at 65 dB SPL by calibrating through the headphones using a Larson Davis 824 sound-level meter and an AEC201 coupler. Total trial duration was 3.25 sec. Starting SNR was –4 dB, about 10-15 dB above the expected SRTs for normal hearing listeners. SRT was calculated as the average SNR of the final 19 of 25 total trials. SRTs were estimated twice for each listener to enable calculation of test-retest reliability coefficients. Reproducibility of the results can also be affected by learning/practice effects. However, the practice effect is expected to be very small when a closed set of small and simple speech stimuli (e.g., digits) is used (Smits et al. 2013; Vlaming et al. 2014). The order of DIN testing and retesting was as follows: BB, 2 kHz, 4 kHz, 8 kHz, BB, 2 kHz, 4 kHz, 8 kHz. Each test block took about 15 minutes to deliver,

### Analysis

R software (version 3.4.2) was used for statistical computing and graphics. DIN-SRT and PTA data were analyzed and compared using Pearson correlation statistics. Receiver operating characteristic (ROC) curves based on these data were plotted. A measure of overall accuracy and sensitivity of a test is the area under the ROC curve. Larger values of the ROC area indicate, on average, more accurate screening tests. One-way analysis of variance (ANOVA) and Tukey post-hoc tests were also used for group comparisons. All *p*-values were two-sided, and a *p*-value of < 0.05 was the statistical significance level.

## RESULTS

Results reported previously from normal hearing (NH) listeners (n=60; Motlagh Zadeh et al. 2019) showed that, as noise bandwidth broadened, mean SRT increased as expected, due to increased masking of higher frequency components of the digits (Figure 3, open symbols). Note that even the broadest low-pass filter (8 kHz cutoff) produced a mean SRT significantly better (3.2 dB more negative; *p* < 0.0001) than that resulting from the unfiltered, broadband noise. This finding showed that sound energy above the upper frequency limit of the standard audiogram (8 kHz) contributes significantly to intelligibility of the digits. These data from NH listeners form the comparative background for the remainder of this report on listeners with mild HI.

**Figure 3.**
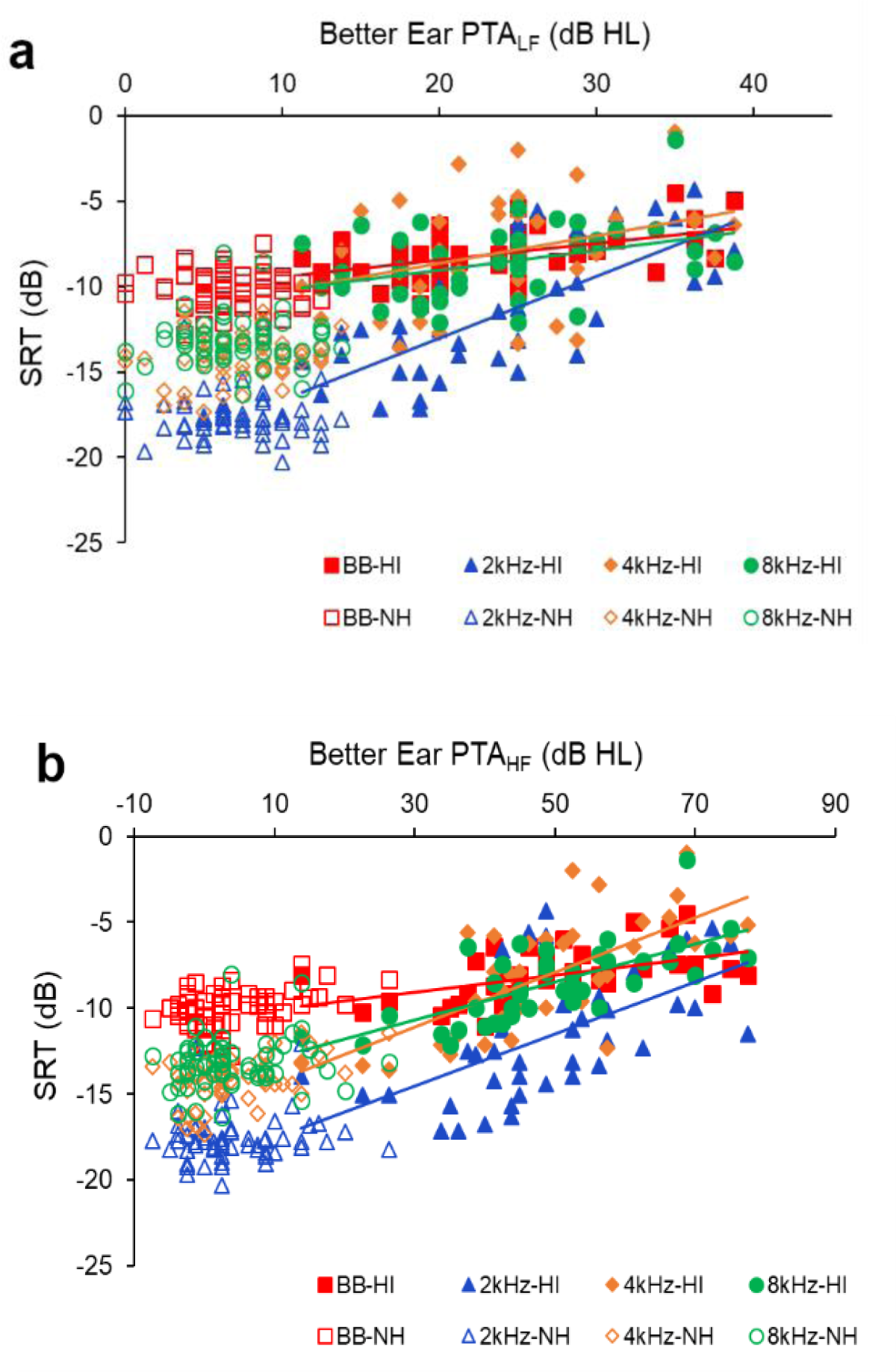
Speech reception thresholds (SRTs) as a function of better ear (Panel **a**) PTA_LF_ and (Panel **b**) PTA_HF_ in normal hearing (NH, open symbols) and hearing impaired (HI, filled symbols and least-square regression lines) groups. LF and HF ranges are defined in Fig. 2. Note that HI was defined by reference to standard audiometry (0.25-8 kHz) throughout, so some HI listeners had some PTA_LF_ thresholds < 20 dB HL and some NH listeners had some PTA_HF_ thresholds > 20 dB HL. A more negative SRT means better hearing. **BB:** Broadband noise, **2 kHz:** 2 kHz low-pass filter noise, **4 kHz:** 4 kHz low-pass filter noise, **8 kHz:** 8 kHz low-pass filter noise.

### Relation to hearing loss

The relation between SRTs with lower-frequency (PTA_LF_) and higher-frequency (PTA_HF_) better ear audiometric averages for the listeners with HI are shown in Figure 3 (filled symbols). The reasons for choosing better ear PTA were, first, the assumption that listeners will preferentially use that ear when they can and, second, typical binaural speech-in-noise tests usually empasize better ear performance (Potgieter et al, 2018; De Sousa et al, 2019).

In contrast to listeners with NH, where SRT stayed relatively flat and invariant across PTA, listeners with HI had an increasing SRT with increasing PTA. Moreover, the relation between an individual’s SRT and PTA became markedly more variable. For example, using the 2 kHz filter, four individuals with mild to moderate hearing loss had SRTs between 5-7dB poorer than the average for that filter (Figure 3b). Conversely, many listeners had SRTs well below the regression line. These and other examples showed that although the SRT-PTA relation was, overall, quite predictable, there were also many exceptions, highlighting the different processes tapped by these two methods. The slopes, intercepts, and correlation of the regression lines for each filter (Figure 3) are given in Table 1. Both better ear PTA_LF_ and PTA_HF_ were significantly correlated with DIN-SRT. SRT for the 2 kHz low-pass filter had the highest correlation with PTA_LF_ (r = 0.71, *P* < 0.0001) and the steepest slope of the regression line. SRTs for the 4 and 8 kHz low-pass filters were most highly correlated with PTA_HF_ (r = 0.70, *P* < 0.0001). However, slopes of the PTA_HF_ regression lines for the three filtered conditions did not differ markedly from each other and were all more than twice as steep as that of the BB condition. Correlations between PTA and SRT were weak and non-significant for NH listeners.

**Table 1.**
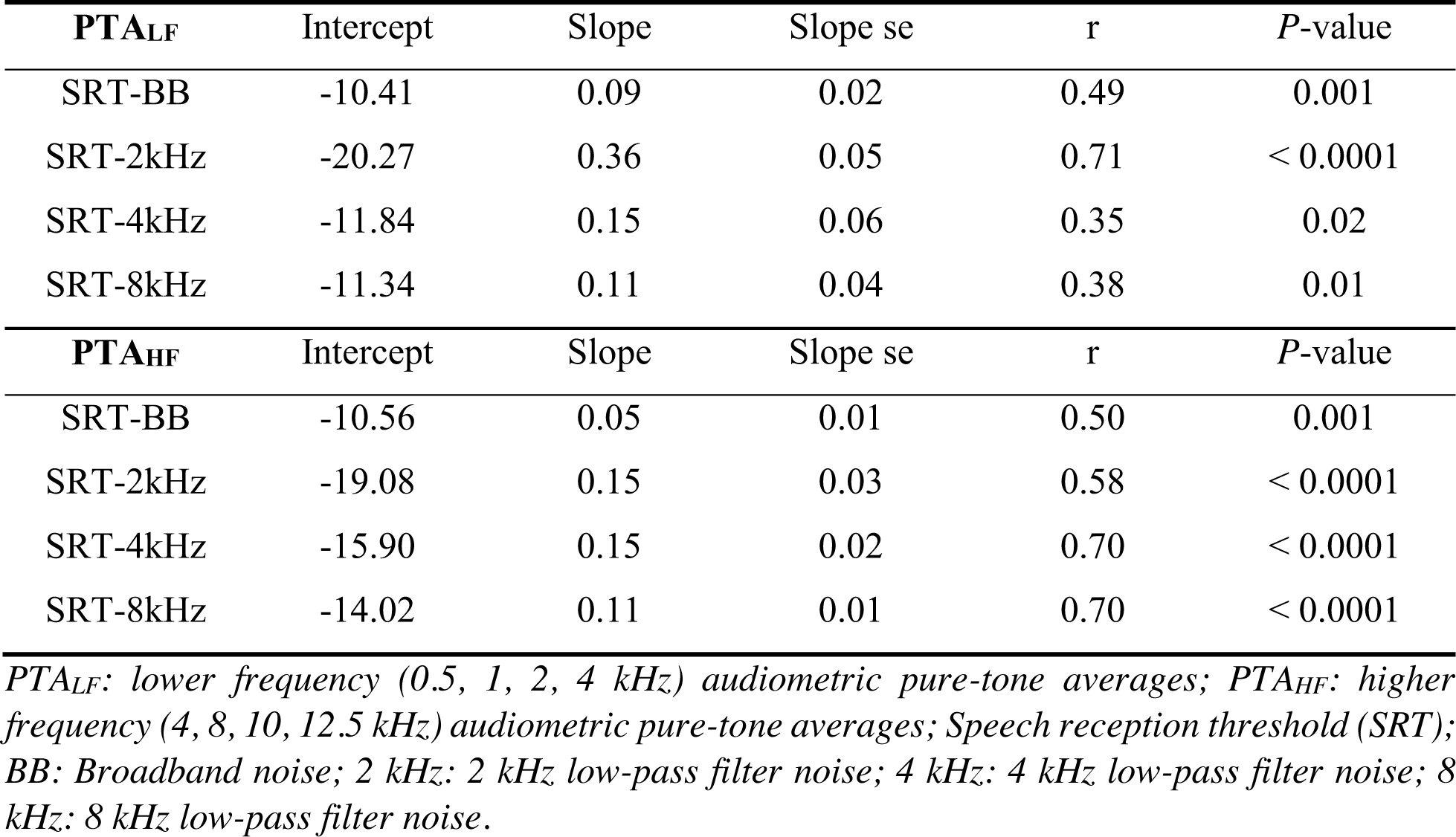
Intercepts (dB), slopes, standard errors of slope, and correlation coefficients (Pearson r) of the regression lines (from Figure 3) for SRT as a function of PTA for hearing impaired listeners.

| **PTA<sub>LF</sub>** | Intercept | Slope | Slope se | $r$ | $P$-value |
| --- | --- | --- | --- | --- | --- |
| SRT-BB | -10.41 | 0.09 | 0.02 | 0.49 | 0.001 |
| SRT-2kHz | -20.27 | 0.36 | 0.05 | 0.71 | < 0.0001 |
| SRT-4kHz | -11.84 | 0.15 | 0.06 | 0.35 | 0.02 |
| SRT-8kHz | -11.34 | 0.11 | 0.04 | 0.38 | 0.01 |
| **PTA<sub>HF</sub>** | Intercept | Slope | Slope se | $r$ | $P$-value |
| SRT-BB | -10.56 | 0.05 | 0.01 | 0.50 | 0.001 |
| SRT-2kHz | -19.08 | 0.15 | 0.03 | 0.58 | < 0.0001 |
| SRT-4kHz | -15.90 | 0.15 | 0.02 | 0.70 | < 0.0001 |
| SRT-8kHz | -14.02 | 0.11 | 0.01 | 0.70 | < 0.0001 |

*PTA<sub>LF</sub>*: lower frequency (0.5, 1, 2, 4 kHz) audiometric pure-tone averages; *PTA<sub>HF</sub>*: higher frequency (4, 8, 10, 12.5 kHz) audiometric pure-tone averages; *Speech reception threshold (SRT)*; *BB*: Broadband noise; *2 kHz*: 2 kHz low-pass filter noise; *4 kHz*: 4 kHz low-pass filter noise; *8 kHz*: 8 kHz low-pass filter noise.

There were also significant correlations between PTA_EHF 10, 12.5, 14, 16 kHz_ (better ear) and DIN SRTs in listeners with EHF hearing loss (>20 dB HL at any frequency from 10-16 kHz; Figure 4). In this analysis, we included all listeners and, for the purpose of regession, assigned them as having an EHF hearing loss irrespective of their lower frequency hearing status. Note that many listeners with normal hearing in the conventional frequency range (open symbols in Figure 4) had an EHF hearing loss. The slopes, intercepts, and correlation of least-square regression lines, fitted to listeners with EHF hearing loss, are shown in Table 2. SRTs of all low-pass filtered noise conditions showed higher correlations with PTA_EHF_ than those of the BB noise condition and, in contrast to the previous PTAs (Table 1), the slope standard error was higher for the BB noise than for the filtered conditions. The 4 kHz low-pass filtered DIN SRT had the highest correlation with PTA_EHF_ (r = 0.81, *P* < 0.0001), and the steepest slope of the regression line with the lowest slope standard error. Correlations between PTA_EHF_ and SRTs were weak and non-significant for listeners with no EHF hearing loss.

**Figure 4.**
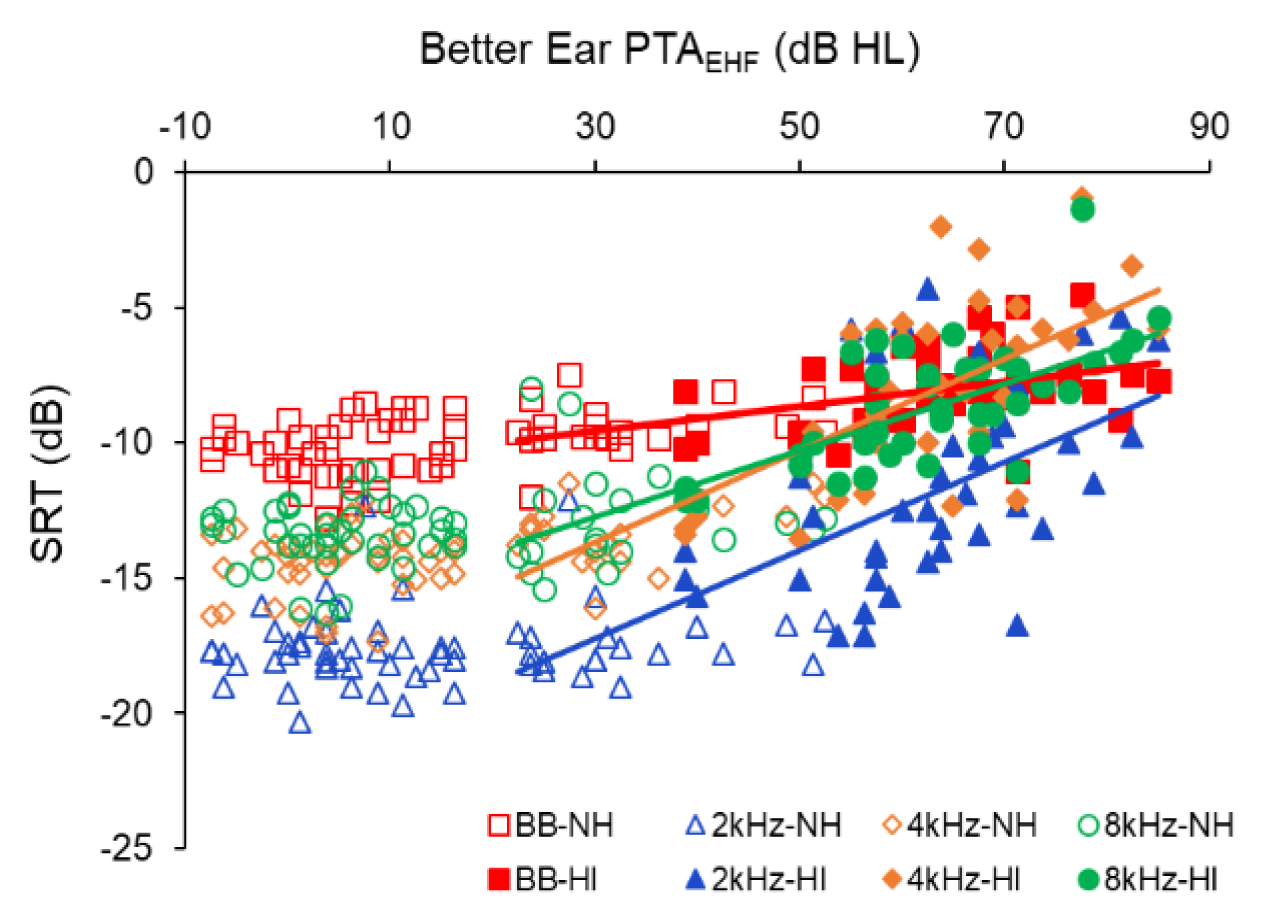
SRTs as a function of better ear PTA_EHF_ (PTA_10,12.5,14,16 kHz_) in normal hearing (NH, open symbols) and hearing impaired (HI, filled symbols) groups. Least-square regression lines were fitted here to listeners with EHF hearing loss, in contrast to Fig. 3, but NH and HI data points remain defined as in previous figures. A more negative SRT means better hearing.

**Table 2.** Intercepts (dB), slopes, standard errors of slope, and correlation coefficients (Pearson r) of the regression lines (from Figure 4) for SRT as a function of EHF-PTA for people with EHF hearing loss.

| $PTA_{EHF}$ | Intercept | Slope | Slope se | $r$ | $P$-value |
| --- | --- | --- | --- | --- | --- |
| SRT-BB | -10.92 | 0.04 | 1.35 | 0.50 | $< 0.0001$ |
| SRT-2kHz | -22.11 | 0.16 | 0.39 | 0.70 | $< 0.0001$ |
| SRT-4kHz | -18.75 | 0.17 | 0.36 | 0.81 | $< 0.0001$ |
| SRT-8kHz | -16.45 | 0.12 | 0.52 | 0.76 | $< 0.0001$ |

*PTA<sub>EHF</sub>: Extended high frequency (10, 12.5, 14, 16 kHz) audiometric pure-tone averages; Speech reception threshold (SRT); BB: Broadband noise; 2 kHz: 2 kHz low-pass filter noise; 4 kHz: 4 kHz low-pass filter noise; 8 kHz: 8 kHz low-pass filter noise.*

To consider further the association between low-pass filtered SRTs and audiometric thresholds, the correlation between SRT of each noise condition with single audiometric frequency (better ear) was calculated for the normal and hearing impaired groups combined (Figure 5). Broadband DIN correlations across the frequency range were weaker than any of the filtered noise conditions, even at lower frequencies. The 2 kHz filtered-DIN test showed a higher correlation for 1 to 4 kHz, while both 4 and 8 kHz filtered noise conditions showed an increased correlation for frequencies ≥ 8 kHz.

**Figure 5.**
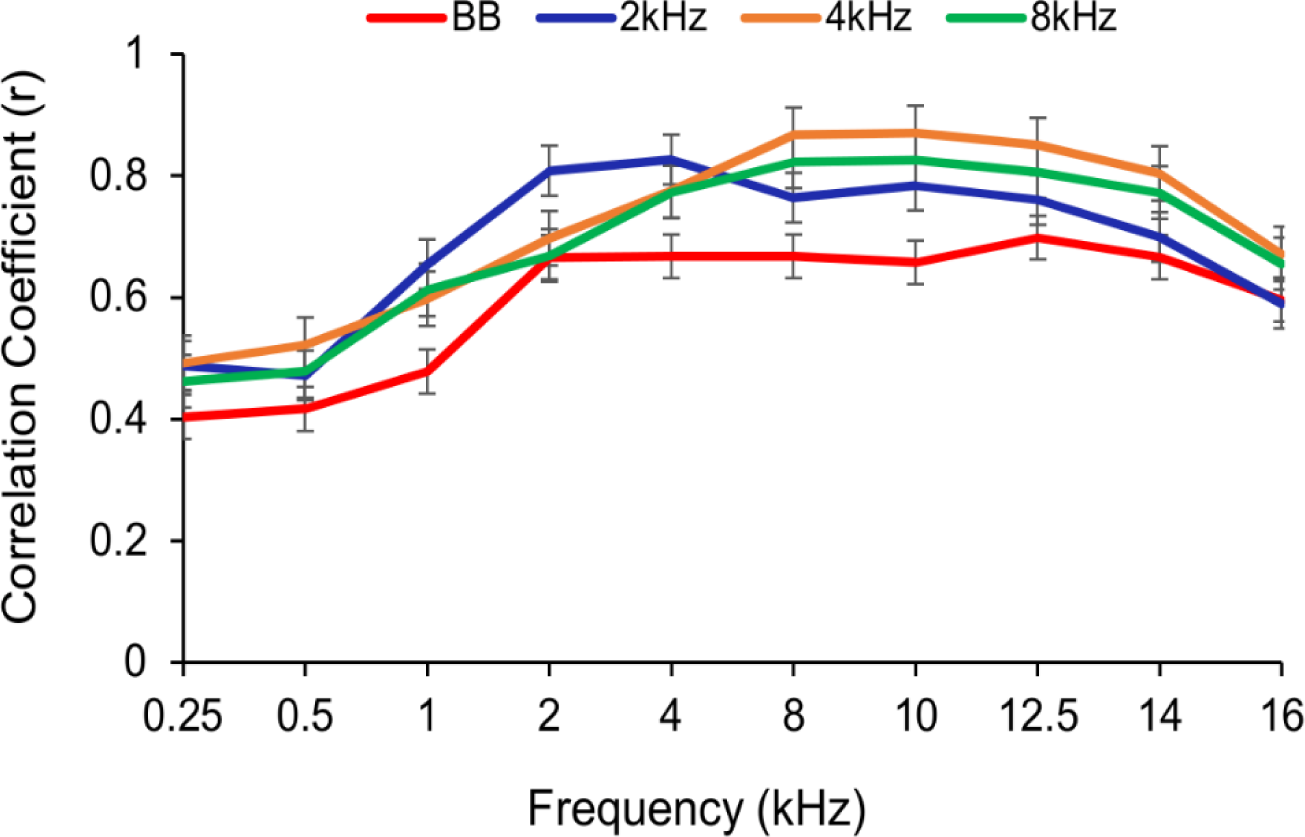
Pearson correlation coefficients (r) between the SRTs of broadband (BB) and low-pass filtered (2, 4, 8 kHz cutoffs) DIN tests with hearing thresholds (better ear) at audiometric and EHF frequencies for the NH and HI groups combined. Error bars are standard errors.

### Sensitivity and Specificity of the DIN tests

Receiver operating characteristics (ROCs) were calculated (Robin et al. 2011) for each test (Figure 6). For this purpose, PTA_HF_ > 20 dB was the criterion for distinguishing normal from impaired high frequency hearing. The cutoff value on the ROC curve classifies the test results as positive or negative. The best cutoff is the threshold (in dB) with the highest sensitivity and specificity. As indicated by black dots in Figure 6, the optimal cutoff value was chosen by taking the point where a further increase of the true positive rate (sensitivity) was equal to an increase of the false positive rate (1-specificity).

**Figure 6.**
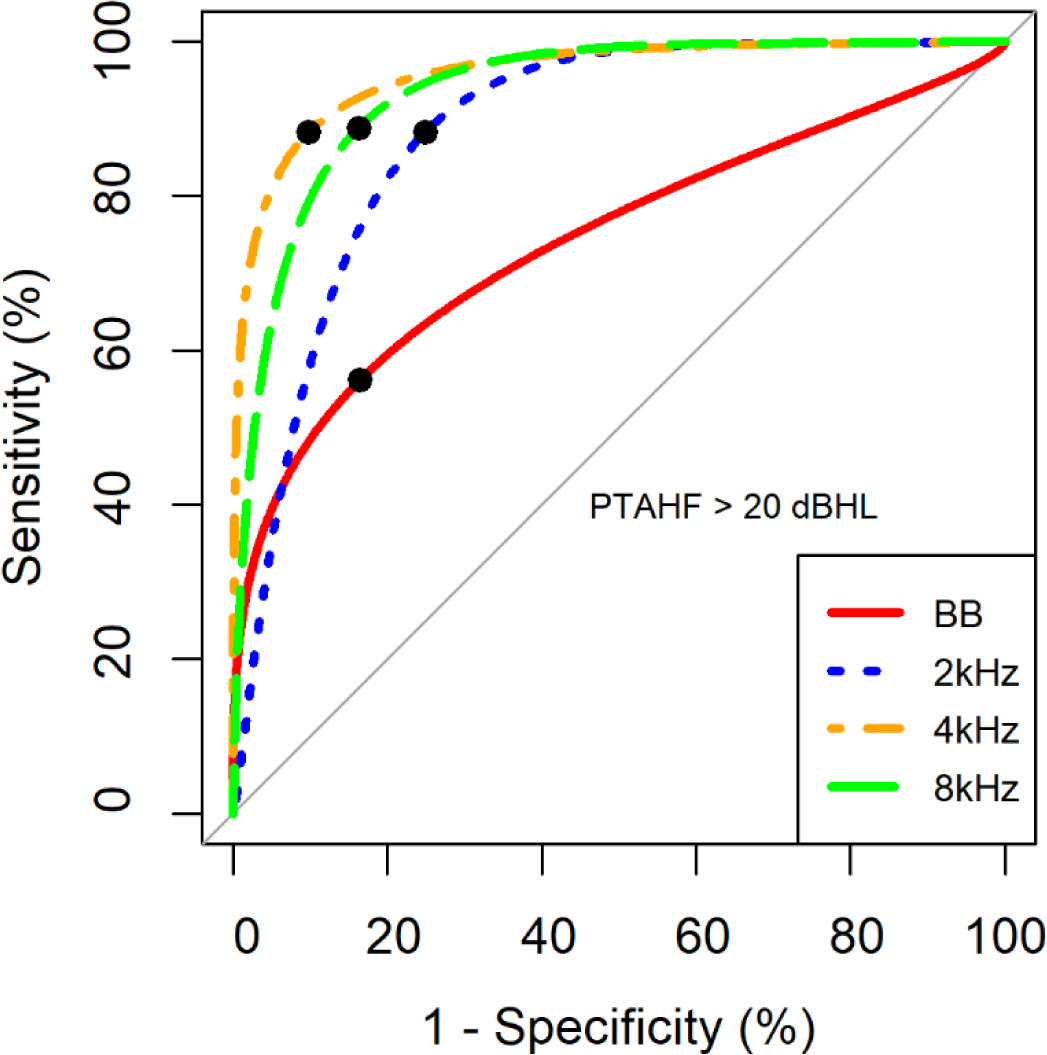
ROC curves showing test characteristics of the broadband (BB) and low-pass filtered DIN test (2, 4, 8 kHz cutoffs) based on PTA_HF_ (4, 8, 10, and 12.5 kHz) for hearing loss of > 20 dB HL. The filled black dots correspond to the chosen optimal cutoff of each test.

Low pass filtered DIN tests had generally higher sensitivity for PTA_HF_ compared to the broadband DIN test (Table 3). Among filtered test conditions, 4 kHz low-pass filtered noise had the maximum ROC area (0.97), and the highest sensitivity (92%). The specificity of 4 and 8 kHz low pass filtered tests were identical (90%). SRTs (optimal cutoff value) were consistent with the regression lines of Figure 3. For lower frequency hearing loss (PTA_LF_ > 20 dB Table 4), 2 kHz low-pass filtered noise masking produced the maximum ROC area (0.90), and the highest specificity (97%). However, sensitivity of 2 kHz noise filtering was lower than the other noise conditions.

**Table 3.** ROC area, sensitivity and specificity of tests, SRT values (optimal cutoffs) obtained from ROC curves for PTA_HF_ > 20 dB HL.

| Test | ROC area | Sensitivity (%) | Specificity (%) | SRT (dB) |
| --- | --- | --- | --- | --- |
| BB | 0.76 | 70 | 85 | -8.6 |
| 2 kHz | 0.90 | 82 | 85 | -15.2 |
| 4 kHz | 0.97 | 92 | 90 | -12.5 |
| 8 kHz | 0.94 | 90 | 90 | -11.3 |

*ROC, receiver operating characteristic; SRT, speech reception threshold; BB: Broadband noise; 2 kHz: 2 kHz low-pass filter noise; 4 kHz: 4 kHz low-pass filter noise; 8 kHz: 8 kHz low-pass filter noise; $PTA_{HF}$ : higher frequency (4, 8, 10, 12.5 kHz) audiometric pure-tone averages.*

**Table 4.** ROC area, sensitivity and specificity of tests, SRT values (optimal cutoffs) obtained from ROC curves for PTA_LF_ > 20 dB HL.

| Test | ROC area | Sensitivity (%) | Specificity (%) | SRT (dB) |
| --- | --- | --- | --- | --- |
| BB | 0.80 | 84 | 71 | -8.6 |
| 2 kHz | 0.90 | 72 | 97 | -12 |
| 4 kHz | 0.85 | 80 | 77 | -9.7 |
| 8 kHz | 0.81 | 84 | 71 | -10.2 |

*ROC, receiver operating characteristic; SRT, speech reception threshold; BB: Broadband noise; 2 kHz: 2 kHz low-pass filter noise; 4 kHz: 4 kHz low-pass filter noise; 8 kHz: 8 kHz low-pass filter noise; $PTA_{LF}$ : lower frequency (0.5, 1, 2, 4 kHz) audiometric pure-tone averages.*

### Test–retest reliability

To enable calculation of test-retest reliability and learning effects, SRTs were estimated twice for each listener (Figure 7). High and significant two-way mixed single intraclass correlation coefficients (ICCs) indicated high levels of internal consistency and reliability of the tests (Table 5). Slopes of the regression lines close to unity (≥ 0.9) and small T1 – T2 values demonstrated small learning effects.

**Figure 7.**
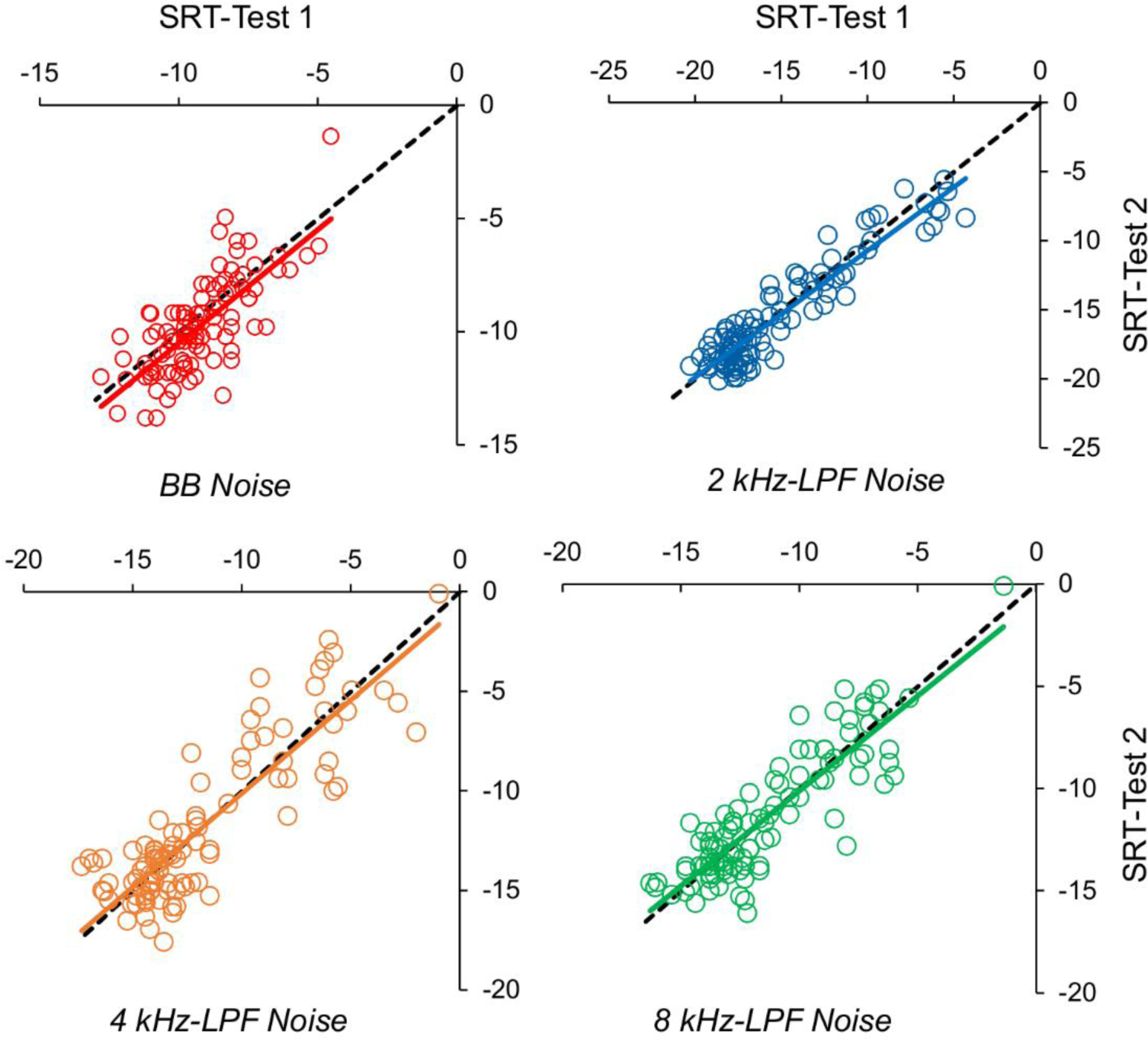
Test-retest SRTs for each noise type in normal and hearing impaired groups combined. Solid colored lines show best fit regressions and dashed black lines show Test 1 = Test 2.

**Table 5.** Digits-in-noise test-retest. Regression line intercepts, slopes, standard errors of slope, intraclass correlation coefficient (ICC), and test-retest mean SRT differences (T1-T2) for the normal and hearing impaired groups combined (from Figure 7).

|  | Intercept | Slope | Slope se | ICC | *P*-value | T1-T2 (dB) |
| --- | --- | --- | --- | --- | --- | --- |
| BB | -0.48 | 1.00 | 0.09 | 0.70 | < 0.0001 | 0.49 |
| 2 kHz | -1.54 | 0.91 | 0.03 | 0.93 | < 0.0001 | 0.27 |
| 4 kHz | -0.76 | 0.93 | 0.05 | 0.80 | < 0.0001 | 0.01 |
| 8 kHz | -0.80 | 0.93 | 0.05 | 0.87 | < 0.0001 | 0.02 |

*SRT, speech reception threshold; BB: broadband noise; 2 kHz: 2 kHz low-pass filter noise; 4 kHz: 4 kHz low-pass filter noise; 8 kHz: 8 kHz low-pass filter noise; T1-T2: mean SRT of Test1 minus that of Test2.*

## DISCUSSION

Low-pass filtered noise increased sensitivity of the DIN for detection of HFHL relative to the standard broadband noise. Of the filters used, DIN sensitivity to higher frequency hearing loss was greatest using the 4 kHz low-pass filter. These results suggest that low-pass filtered noise may be usefully substituted for broadband noise to improve earlier detection of HFHL using DIN test.

### Relation to HFHL

In the hearing impaired group, the relation between PTA_LF_ and DIN-SRTs showed that the 2 kHz low-pass filter was the best noise condition for detecting hearing loss between 0.5 to 4 kHz. For PTA_HF_, the 2 kHz low-pass filter was more sensitive than broadband noise to HFHL (4-12.5 kHz), but not as sensitive as the 4 and 8 kHz filters. Vlaming et al (2014) used a single, 1.5 kHz low-pass filtered noise masker and compared those SRTs with broadband masked SRTs for PTA_LF_ (0.5, 1, 2, and 4 kHz) and PTA_HF_ (3, 4, 6, and 8 kHz). Consistent with findings reported here (2 kHz filtered noise), they found a steep regression slope and high correlation between the PTA_HF_ and SRTs for the filtered noise (r = 0.79) compared to broadband noise (r = 0.62). However, in contrast to our findings, PTA_LF_ in their study was less correlated with the SRTs of the filtered noise (r = 0.66) than the SRTs of the broadband noise (r = 0.72). The reason for this discrepancy may be because three listeners in their hearing loss group had better SRTs (between −20 to −25 dB) in the 1.5 kHz filtered noise condition, resulting in lower correlation for the filtered, than for the broadband noise. More generally, the group of participants with HI studied here had more homogeneous PTAs than participants with HI studied by Vlaming et al. (2014), possibly explaining the observed inconsistency. They argued that high correlation of filtered noise SRTs with PTA_HF_ and medium correlation with PTA_LF_ demonstrated increased sensitivity of the filtered-noise DIN tests for high-frequency (3–8 kHz) hearing loss. However, we found that 2 kHz filtering also improved sensitivity of the DIN to hearing loss below 4 kHz, relative to broadband filtering.

The correlation, sensitivity and specificity of the 2 kHz low-pass filter for detecting HI across the spectrum were also high, suggesting this (or the 1.5 kHz filter used previously; Vlaming et al. 2014) could be a more useful filter to choose for detecting hearing loss between 0.25-4 kHz. This finding is also consistent with the findings of Jansen et al. (2014) and Leensen et al. (2011) that showed low-pass filtering of broadband noise at 1.4 kHz improves sensitivity of the Dutch CVC-words-in-noise task to detect HFHL. In contrast to the findings of the present and the aforementioned studies, Vercammen et al (2018) reported no significant benefit of low-pass filtering of the masker (cutoff at 1.5 kHz) in a Flemish DIN task over the broadband noise masker in terms of area under the ROC curve. This appeared to be because the study of Vercammen et al. (2018) included older people in the normal hearing group, set a higher criterion for hearing impairment (PTA_1, 2, 4, 8 kHz_ > 25 dB HL), and adjusted for age in the calculations of sensitivity and specificity for the low-pass masked thresholds. Athough the age adjustment was appropriately justified on the basis of maintaining comparability with, and maximum prediction of tone thresholds, it seems to us debatable whether PTA rather than DIN-SRT, or either, should have been adjusted as a measure of high frequency hearing *impairment*.

### Relation to EHF hearing loss

In this study, as noise bandwidth broadened in the normal hearing group, mean SRT increased, due to greater masking of the higher-frequency speech components. As shown in our previous report (Motlagh Zadeh et al. 2019), the broadest low-pass filter (8 kHz cutoff) produced a better SRT than that resulting from the broadband noise, revealing the sensitivity of the DIN test to EHF (10-16 kHz) cues. Here, we extended the frequency range for PTA_HF_ up to 12.5 kHz to examine the sensitivity of the DIN tests to higher frequency HFHLs that are very common, even in otherwise normally hearing young adults (Motlagh Zadeh et al. 2019). Our results showed highest correlations between DIN-SRT and PTA_HF_ for the 4 and 8 kHz filtered noise conditions in the hearing-impaired group, supported by high sensitivity (≥ 90%) and specificity (90%) for detecting an average HFHL of 20 dB or more. We also found strong, significant correlations between PTA_EHF_ and DIN-SRT in listeners with EHF hearing loss, revealing the potential of EHF hearing thresholds in predicting speech perception in noise performance. Sensitivity of the 4 kHz filtered noise and its correlation with both PTA_HF_ and PTA_EHF_ were relatively better than the 8 kHz filter noise. For PTA_HF_, this may be because of the presence of additional significant signal energy between 4-8 kHz when using the 4 kHz low-pass filtered noise. For PTA_EHF_, an explanation is not as clear. It is likely that individuals with impaired hearing in the EHF range also have some degree of impaired hearing in the range 4-8 kHz, resulting in higher SRTs and, hence, higher sensitivity of the test than listeners who lack EHF hearing loss. To test this possibilty we compared PTA of 4 and 8 kHz for participants with and without PTA_EHF_ hearing loss. A significant relationship was found between PTA_4-8 kHz_ and PTA_EHF_ (*P*-value < 0.0001, F_1, 98_ = 95.94, r = 0.88).

### Clinical Application

Even though sensitive hearing screening in the conventional frequency range is necessary for timely diagnosis of hearing loss, it may not be sufficient for detection of early signs of hearing loss when preventive methods could be effectively deployed. Optimizing a screening method with sensitivity to hearing loss at extended high frequencies (> 8 kHz) could lead to prevention and follow-up of potentially more disabling hearing loss in the lower frequency range of hearing later in life. The audibility of very high frequency tones is most sensitive in young children and it becomes progressively less sensitive throughout the remainder of life (Rodríguez Valiente et al, 2014). For example, a downward decline for hearing 20 kHz starts from 4-6 years of age (Rodríguez Valiente et al, 2014; Trehub et al, 1988). If the criterion for hearing loss is extended to the EHF range of hearing, many young adults (or even children) might be considered as candidates for active monitoring, protection, and some form of hearing intervention to prevent subsequent disabling hearing loss (presbycusis) at lower frequencies which are most vital for speech perception. The strong, significant correlations of 4 and 8 kHz low-pass filtered DIN-SRT with PTA_HF_ and PTA_EHF_ suggest the advantage of these higher frequency filters for screening and confirmation of early detection of HFHL. The low-pass filtered DIN test also has the potential to be developed as a sound field test to measure supra-threshold speech recognition ability in more realistic listening conditions than a headphone-delivered test, and as a convenient way for testing children and adults while using their hearing devices (e.g., hearing aids and cochlear implant).

Recently, Moore and colleagues (2019) developed a DIN test that uses sound-field presentation of digits against a multi-talker babble masker (“FreeHear”). The masker is either co-located with the digits (0°), or presented 90° either side of the participant, enabling measurement of spatial ‘release from masking’, the benefit received from a spatial mis-match between target and masker. Based on the current study, a future direction of FreeHear may involve adding low-pass filtering (e.g., cutoff at 4 kHz) to the babble-masker, or to a conventional speech shaped noise within the sound-field DIN, to sensitize that test to EHF hearing loss and to examine the role of EHF hearing in a more realistic listening environment. The findings of this study support the development of simple, self-administered ways to screen for HFHL, and next-generation hearing aids with a higher frequency response range.

## Data Availability

The data that support the findings of this study are available on request from the corresponding author.

## ACKNOWLEDGEMENTS

This study was supported by NIH grant R21DC016241 and by the Cincinnati Children’s Hospital Research Foundation. David Moore receives support from the NIHR Manchester Biomedical Research Centre. L.M.Z., N.H.S., D.W.S and D.R.M. designed the experiments. L.M.Z. collected the data. L.M.Z. and D.R.M analyzed the data and wrote the manuscript.

